# A recombinant BCG-based vaccine against the human respiratory syncytial virus induces a balanced cellular immune response against viral and mycobacterial antigens

**DOI:** 10.1101/2022.02.07.22270648

**Authors:** Gaspar A. Pacheco, Nicolás M. S. Gálvez, Catalina A. Andrade, Yaneisi Vázquez, Linmar Rodríguez-Guilarte, Pablo A. González, Susan M. Bueno, Alexis M. Kalergis

## Abstract

**BACKGROUND:** The human respiratory syncytial virus (hRSV) is a respiratory pathogen responsible for most cases of acute lower respiratory tract infections in infants worldwide. Although this virus represents a significant social and economic burden, there are no safe and effective available vaccines. rBCG-N-hRSV is a vaccine candidate consisting of a recombinant attenuated *Mycobacterium bovis* Bacillus Calmette-Guérin (BCG) expressing the nucleoprotein of hRSV (N-hRSV).

**METHODS:** rBCG-N-hRSV was applied intradermally in three different doses (5×10^3^, 5×10^4^, or 1×10^5^ CFU) to healthy adults enrolled in a randomized, double-blind, dose-escalating phase 1 clinical trial (NCT03213405). Blood samples were taken before and at various time points after immunization. Cellular and humoral immune parameters were assessed by analyzing circulating immune cells and sera, respectively.

**RESULTS:** Perforin- and Granzyme B-producing PBMCs recognizing viral or mycobacterial antigens were found to increase after immunization with rBCG-N-hRSV. These cells also upregulated IFN-γ and IL-10 secretion in response to N-hRSV and upregulated IFN-γ, IL-6, and TNF-α secretion in response to mycobacterial proteins. While naïve T cell populations contracted over time, no specific memory T cell subset expanded significantly. Although binding to C1q by anti-N-hRSV or anti-mycobacterial antibodies decreased slightly after immunization, no apparent changes were found in the concentration of IgG subclasses against N-hRSV or mycobacterial antigens.

**CONCLUSIONS:** The immune response elicited by immunization with rBCG-N-hRSV consists mainly of antigen-specific T cells. The data reported here provide novel information about the characteristics of the immune response elicited after immunization with rBCG-N-hRSV, supporting the safety and immunogenicity of this vaccine.

## INTRODUCTION

The human respiratory syncytial virus (hRSV) is the leading etiological agent for acute lower respiratory tract infections (ALRTISs) in children under 5 years old, the elderly, and immunocompromised patients [1]. This virus is responsible for over 30 million hospitalizations and over 200,000 deaths per year worldwide [1], [2]. Typical symptoms of ALRTIs include nasal congestion, fever, wheezing, bronchoconstriction, bronchial and alveolar collapse due to excessive mucus production, and pneumonia [3]. However, symptoms associated with hRSV infection may reach far beyond acute disease, as infection with hRSV may lead to sequelae, such as the development of asthma later in life and neurological and cognitive impairment [4]–[6]. Despite the substantial social and economic burden that hRSV poses, there are no safe and effective vaccines against this virus approved to date. One of the current vaccine candidates against hRSV is rBCG-N-hRSV, a vaccine based on an attenuated *Mycobacterium bovis* Bacillus Calmette-Guérin (BCG) that recombinantly expresses the nucleoprotein of hRSV (N-hRSV) [7]– [11]. The design of this vaccine is highly advantageous as BCG is one of the oldest and safest vaccines used to date and is currently applied to newborns to prevent tuberculosis in most countries [12], [13]. Thus, this BCG strain could potentially grant protection against both tuberculosis and hRSV in newborns, which represent the most at-risk population of hRSV-caused ALRTIs. rBCG-N-hRSV has been thoroughly tested in animal models and has shown to induce protective humoral and cellular immunity against hRSV after viral challenge in both, mice and calves [8]–[11]. Importantly, a randomized, double-blind, dose-escalating phase 1 clinical trial was carried out to evaluate the safety, tolerability, and transmissibility of this live attenuated vaccine candidate in healthy male adults [7]. The vaccine was found to be safe and well-tolerated in adults at all tested doses, and no evidence of transmissibility of this BCG strain was found. Moreover, preliminary immunogenicity data were generated, finding that rBCG-N-hRSV induces a cellular immune response characterized by enhanced IFN-γ and IL-2 secretion after stimulation with either N-hRSV or mycobacterial antigens, especially at the higher doses tested [7]. The primary cell type associated with the secretion of these cytokines was presumed to be CD4^+^ helper T cells. Total IgG against N-hRSV or mycobacterial antigens were not found to be statistically increased after immunization. Here, we provide new data and a more in-depth characterization of the cellular and humoral immune response elicited by immunization with the rBCG-N-hRSV vaccine.

## METHODS

### Phase 1 Clinical Trial study design and sample collection

A randomized, double-blind, dose-escalating Phase 1 Clinical Trial was carried out (NCT03213405), which primary and secondary outcomes evaluated were safety, tolerability, transmissibility, and immunogenicity of the rBCG-N-hRSV vaccine in healthy adults [7]. The study protocol followed current ethical guidelines, such as Tripartite Guidelines for Good Clinical Practices, the Declaration of Helsinki [14], and local regulations. This study was approved by both, the Institutional Ethical Committee (number 15216) and the Institute of Public Health of Chile (ISP Chile, number EC819077/16). Inclusion and exclusion criteria and additional information about this clinical trial was reported previously [7]. Briefly, 24 male adults aged 18-50 years were enrolled and randomized into three cohorts consisting of eight subjects each. Two participants were immunized intradermally in the deltoid area with 2×10^5^ CFU of BCG-WT in each cohort [7]. The remaining six subjects were vaccinated with escalating doses of rBCG-N-hRSV (5×10^3^, 5×10^4^, or 1×10^5^ CFU) [7]. Data obtained from subjects immunized with BCG-WT were analyzed together, independently of their cohort [7]. Blood samples were taken before immunization (Day 0) and 14, 30, 60, 120, and 180 days post-immunization [7]. Peripheral blood mononuclear cells (PBMCs) and sera were isolated from these blood samples and stored in liquid nitrogen and in a -80°C freezer, respectively.

### Perforin/Granzyme B ELISPOT Assays

The secretion of Perforin (Perf) and Granzyme B (GrzB) by PBMCs was evaluated using the Human Granzyme B/Perforin Double-Color Enzymatic ELISPOT Assay (ImmunoSpot). The assay was performed according to the manufacturer instructions, incubating the cells with each stimulus for 48 hours at 37°C and 5% CO_2_. 2×10^5^ PBMCs were plated and stimulated with either 1.25 µg/mL N-hRSV (Genscript) or 750 IU/mL bovine PPD (ThermoScientific). Positive and negative stimulation controls were also included as internal controls (data not shown). After developing the plates, these were air-dried face-down on a paper towel for 24 h, and then stored with the underdrain on until spots were counted with an ImmunoSpot S6 CORE Analyzer (Immunospot).

### Serum cytokine measurements by Cytometric Bead Array^™^

Cytokine measurements were performed using the BD^™^ Cytometric Bead Array (CBA) Human Th1/Th2/Th17 Cytokine Kit (BD Biosciences). Determinations were performed following the manufacturer instructions, with either undiluted sera or culture supernatant of PBMCs diluted 1:4. The median fluorescent intensity of PE fluorochrome (MeFI) in each bead population was calculated for every sample. A 4-parameter logistic (4PL) standard curve was constructed using PE MeFI for each cytokine, subtracting the signal from the blank tube to each sample and standard. Cytokine concentrations were obtained from the respective standard curve.

### Memory T cell quantification by flow cytometry

PBMCs stored in liquid nitrogen were thawed, stimulated with the indicated antigens, and analyzed as previously described [7]. Briefly, cells were pelleted, resuspended, and stimulated with 2.5 µg/mL N-hRSV or 25 µg/mL PPD for 5 hours and then incubated for 16 hours with a cocktail of secretion inhibitors [7]. Staining was performed as previously described [7] and data were acquired in a BD LSRFortessa X-20^™^. Gating and analyses were performed using the FlowJo software (V10.6.2).

### C1q-binding assays

C1q binding to antibodies was measured by ELISA on plates coated O.N. at 4°C with 50 µL of either 1 µg/mL N-hRSV (Genscript) or 25 µg/mL bovine PPD (ThermoScientific) in 100 mM bicarbonate/carbonate pH 9.5 buffer. Then, plates were washed three times with PBS-Tween 20 0.05% (Wash buffer) and then blocked for two hours with 200 µL of PBS-Milk 5% (Blocking solution). Sera samples were diluted 1:30 in blocking solution and inactivated at 56°C for 30 minutes. Plates were washed three times, incubated with 50 µL of inactivated sera samples for one hour at RT, washed three times, incubated for one hour with 50 µL of 4 µg/mL purified human C1q diluted in blocking solution, washed five times, incubated for one hour with 50 µL of mouse anti-human C1q (Invitrogen) diluted 1:2,000, washed seven times, incubated for two hours with 50 µL of rat anti-mouse IgG1-HRP (Invitrogen) diluted 1:2,000, washed seven times with 200 µL of wash buffer and then washed manually one time with 200 µL of PBS. 50 µL of TMB (ThermoScientific) were added and incubated for 15 minutes at RT. Then, 50 µL of 2 N sulfuric acid were added and the absorbance at 450 nm was immediately measured. A C1q standard curve was incorporated in every plate. A C1q Binding Index was defined and calculated as follows:

C1q Binding Index = Bound C1q (ng) / Log_10_ (Antibody Concentration [ng/mL])

### IgG subclass quantification by ELISA

ELISA plates were coated O.N. at 4°C with 50 µL of 2 µg/mL N-hRSV (Genscript) or 25 µg/mL bovine PPD (ThermoScientific) in sodium carbonate/bicarbonate 100 mM pH 9.5 buffer. Then, plates were washed three times with 200 µL of PBS-Tween 20 0.05% (Wash buffer). Plates were blocked with PBS-FBS 10% (Blocking solution) for two hours at RT, washed three times, incubated with sera diluted 1:5 in blocking solution for two hours, washed three times, incubated for one hour with 50 µL of either rat anti-human IgG1 (BioLegend) or rat anti-human IgG2 (BioLegend) diluted 1:500, washed three times, incubated for one hour with 50 µL of rat anti-mouse IgG1-HRP (Invitrogen) diluted 1:2,000, washed three times with 200 µL of wash buffer and manually washed one time with 200 µL of PBS. 50 µL of TMB Substrate Reagent (ThermoScientific) were added to each well. 50 µL of 2 N sulfuric acid were added to each well and absorbance at 450 nm was immediately measured. Standard curves for IgG1 or IgG2 were incorporated in every plate.

### Statistical analyses

All graphics and statistical analyses were performed in GraphPad Prism, version 9.1.0. Analyses of the standard curves obtained in ELISA assays were performed by an unconstrained 4-parameter logistic regression (4PL). Data were then interpolated in those regressions. To calculate fold-changes relative to pre-immune conditions, the observed response variable at a given time was divided by that observed at Day 0. A two-way ANOVA for repeated measures with *post-hoc* Dunnet’s test corrected for multiple comparisons against Day 0 was performed to evaluate differences after immunization. Base 10 logarithms of fold changes were calculated before statistical analyses.

## RESULTS

### A single dose of rBCG-N-hRSV induces moderate inflammation in healthy adults

The safety and reactogenicity of three escalating doses of rBCG-N-hRSV had been studied previously [7]. However, molecular immune parameters that support that this vaccine does not induce systemic hyper-inflammation had not been measured. Thus, we evaluated the concentration of various cytokines in sera of immunized subjects. Cytokine concentrations was determined at days 0, 14, 30, 60, 120, and 180 post-immunization for subjects immunized either with one of the three doses tested of rBCG-N-hRSV, or a standard dose of BCG-WT (**Fig. 1** and **Supp. Fig. 1**). Cytokine responses were heterogenous among subjects within the same group (**Fig. 1**). Nonetheless, all cytokines were found in very low concentrations in the sera, consistent with the lack of an exacerbated inflammation after immunization. Only a slight induction of early IFN-γ was observed in sera from subjects immunized with the highest doses tested for the vaccine (**Fig. 1A**). These results suggest that the application of rBCG-N-hRSV does not induce systemic hyper-inflammation, which further supports the safety of this vaccine candidate in healthy adults.

**Figure 1.**
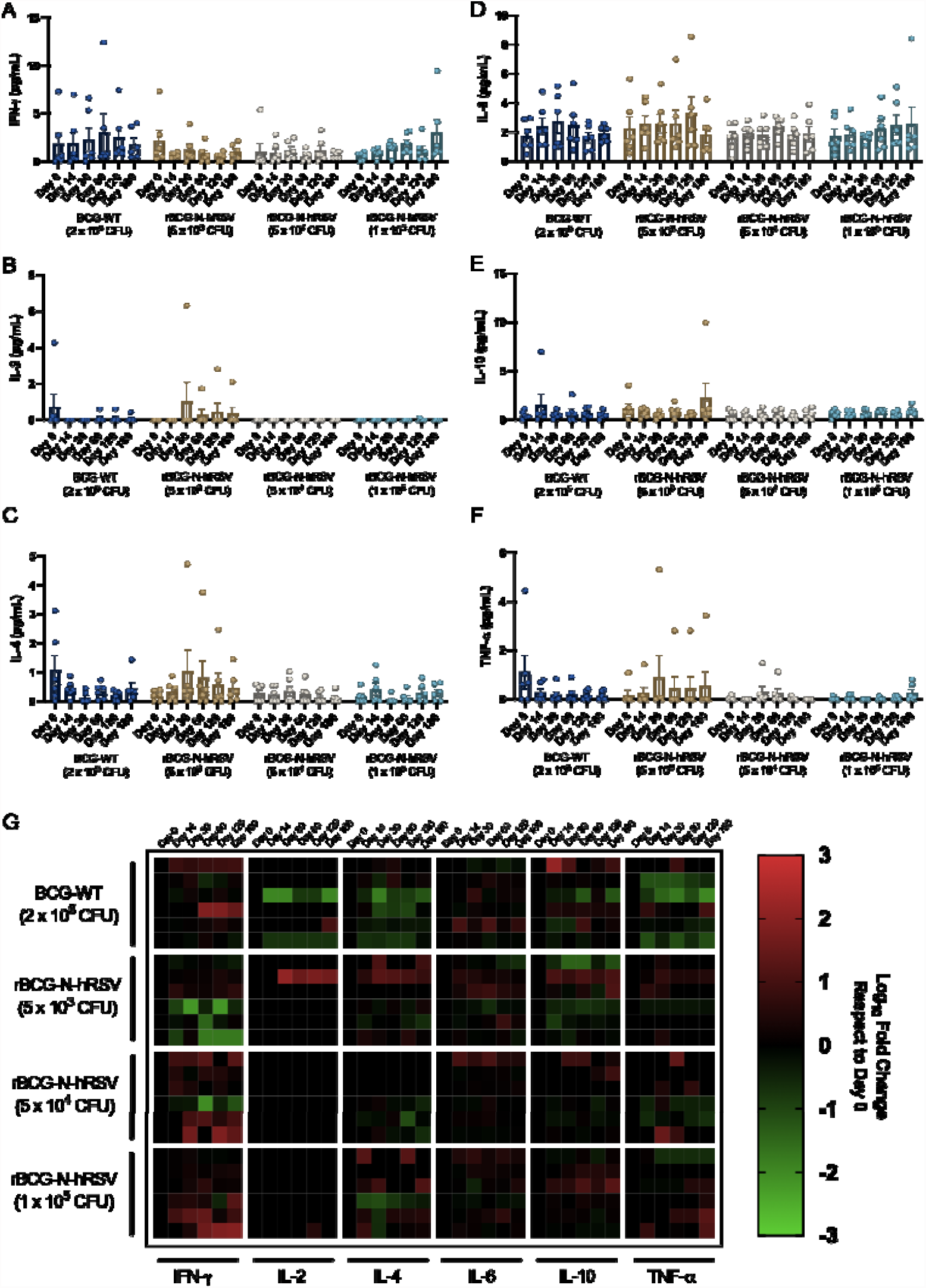
Immunization with rBCG-N-hRSV leads to a moderate systemic cytokine response. The concentration of **(A)** IFN-γ, **(B)** IL-2, **(C)** IL-4, **(D)** IL-6, **(E)** IL-10, and **(F)** TNF-α in sera samples are presented. Bars represent means, and error bars represent SEM. A two-way ANOVA for repeated measures with *post-hoc* Dunnet’s test corrected for multiple comparisons against Day 0 was performed for the analysis of the data. **(G)** Heatmap of log_10_ fold change of the concentration of selected cytokines compared to Day 0. Each block of columns represents a particular cytokine, labeled below. Individual columns represent timepoints after immunization, specified above. Each block of rows represents a particular cohort of immunized study subjects, labeled left. Individual rows represent individual subjects. A color scale is depicted on the right.

### rBCG-N-hRSV primes hRSV- and mycobacterial-specific cytotoxic T cells in healthy adults

One of the advantages of the rBCG-N-hRSV vaccine is the possibility to simultaneously induce, anti-mycobacterial and antiviral immune responses [8]. While the CD4^+^ helper T cell responses against N-hRSV and mycobacterial antigens after rBCG-N-hRSV immunization have already been evaluated [7], the secretion of cytotoxic molecules by CD8^+^ cytotoxic T cells against these antigens had not been previously assessed. The number of Perf^+^ and GrzB^+^ spot-forming cells (SFCs) was counted for each of the PBMCs samples obtained from immunized subjects upon *in vitro* stimulation for 48 h with either N-hRSV (**Fig. 2A-C**) or PPD (**Fig. 2D-F**). Double positive (Perf^+^ GrzB^+^) SFCs were also measured (**Supp. Fig 2**), as well as the fold-changes relative to the pre-immune condition (**Supp. Fig 3**). The stimulation of PBMCs with N-hRSV induced a potent effector cytotoxic molecule response only in subjects immunized with 5×10^4^ CFU of rBCG-N-hRSV (**Fig. 2A-C & Supp. Fig. 3A-C**). A strong Perforin response was observed in PBMCs from subjects of this cohort (**Figure 2B & Supp. Fig. 3A**), which showed a progressive increase in Perforin secretion up to day 60 post-immunization. Granzyme B responses were modest and only slightly increased for these subjects (**Fig. 2C & Supp. Fig. 3B**). PBMCs obtained from individuals immunized with BCG-WT and stimulated with N-hRSV did not produce the evaluated cytotoxic molecules, which was expected. Importantly, the stimulation of PBMCs with PPD (a purified protein fraction of tuberculin) induced an effector cytotoxic response based on molecular markers in subjects immunized with either 2×10^5^ CFU of BCG-WT or 5×10^4^ CFU of rBCG-N-hRSV (**Fig. 2D-F**). As expected, subjects vaccinated with BCG-WT showed a progressive increase in responsiveness against PPD after immunization (**Fig. 2E-F**). Our results suggest that 5×10^4^ CFU of rBCG-N-hRSV is the optimal dose to induce the secretion of cytotoxic molecules by PBMCs in response to either PPD or N-hRSV (**Fig. 2**). Accordingly, the secretion of Perforin and Granzyme B in response to stimulation with N-hRSV is dependent on the expression of N-hRSV by the BCG vaccine. Given that we observed an antigen-specific T cell response in subjects immunized with rBCG-N-hRSV, we sought to determine whether immunization alone could promote the expansion of memory T cell subsets [15] (gating shown in **Supp. Fig. 4**). Stimulation of PBMCs with N-hRSV or PPD led to an apparent contraction for both the naïve CD4^+^ and CD8^+^ T cell populations (**Figs. 3 & 4**). However, neither central memory T cells (T_CM_), effector memory T cells (T_EM_), nor CD45RA-expressing effector memory T cells (T_EMRA_) seemed to expand to a significant extent after one single dose of rBCG-N-hRSV (**Figs. 3 & 4**).

**Figure 2.**
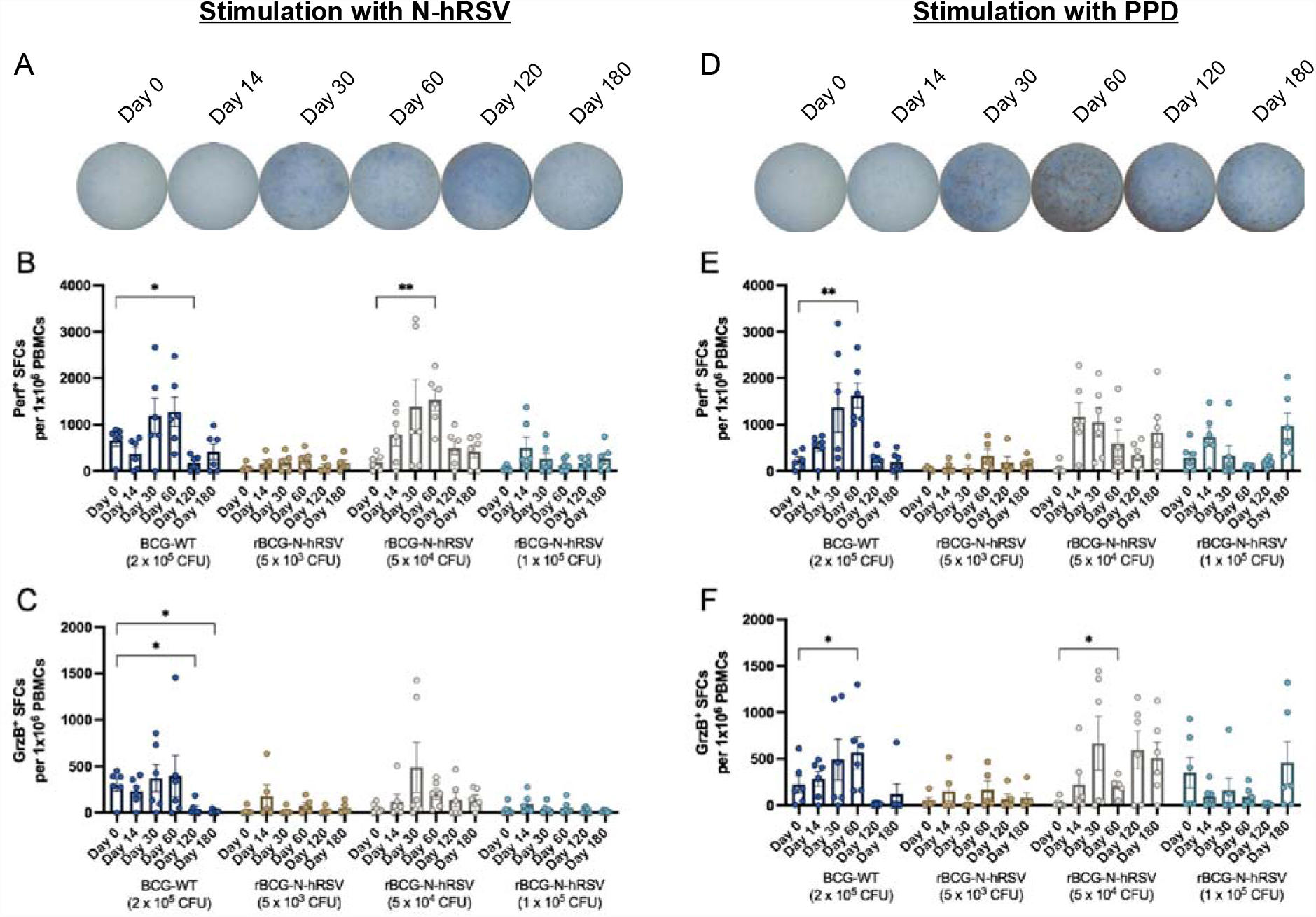
Stimulation with N-hRSV or PPD leads to enhanced Perforin and Granzyme B secretion by PBMCs from subjects immunized with 5×10^4^ CFU of rBCG-N-hRSV. Representative pictures of ELISPOT wells in which PBMCs from subjects immunized with 5×10^4^ CFU of rBCG-N-hRSV were stimulated with either **(A)** N-hRSV or **(C)** PPD. Pictures are representative of subjects immunized with 5×10^4^ CFU of rBCG-N-hRSV. Blue spots are Perf^+^ spot-forming cells (SFCs) and red spots are GrzB^+^ SFCs. **(B & E)** Perf^+^ SFCs and **(C & F)** GrzB^+^ SFCs were counted after PBMCs were stimulated for 48 hours with either 1.25 µg/mL of N-hRSV **(B-C)** or 750 IU/mL of PPD **(E-F)**. Bars represent the mean value of SFCs, and error bars represent the SEM. A two-way ANOVA for repeated measures with *post-hoc* Dunnet’s test corrected for multiple comparisons relative to Day 0 was performed for the analysis of the data. * = P<0.05, ** = P<0.01.

**Figure 3.**
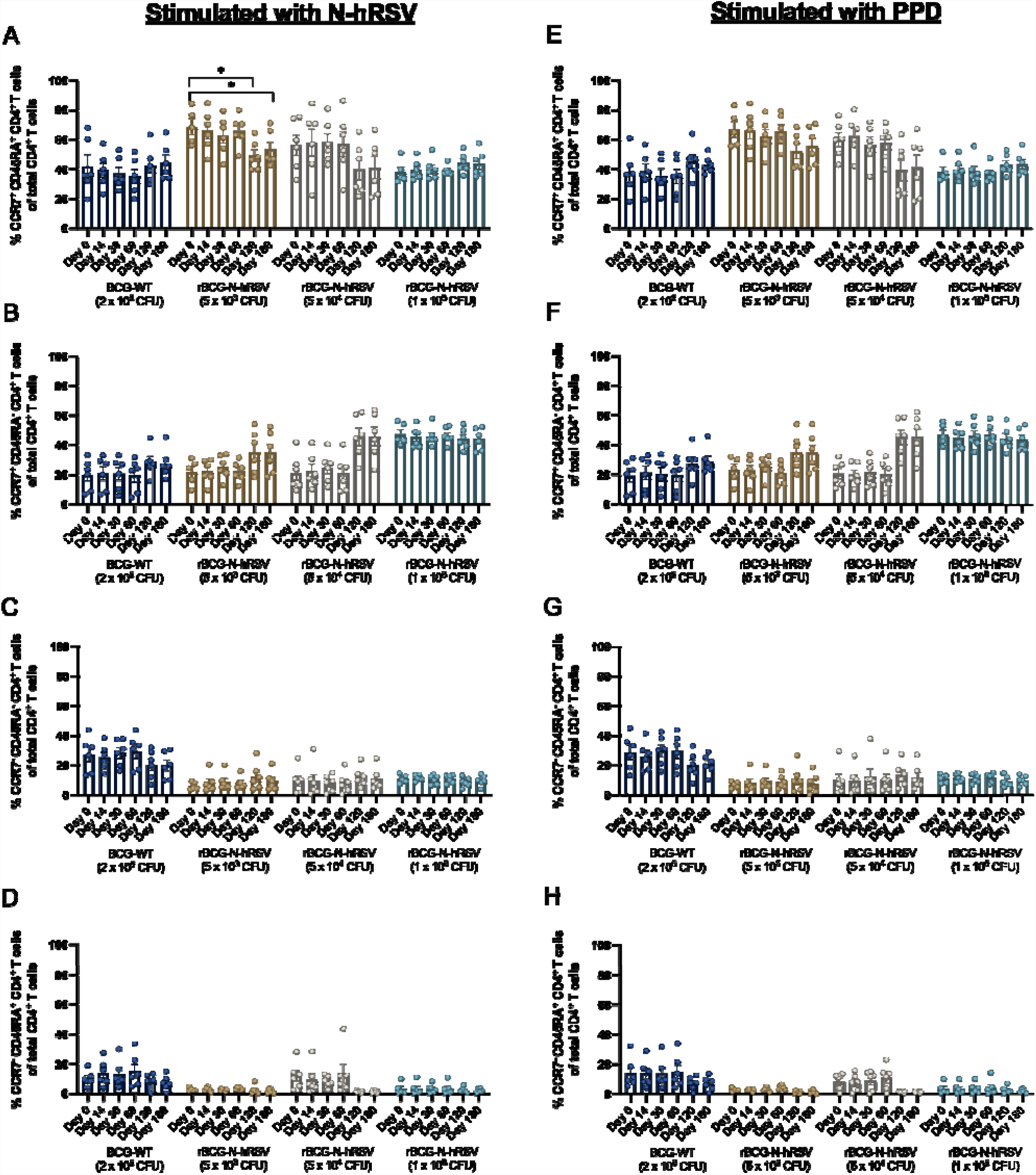
CD4^+^ T cell memory subsets do not expand after a single immunization with rBCG-N-hRSV. T cell subsets defined by the expression of CCR7 and CD45RA were evaluated after stimulation of PBMCs with **(A-D)** N-hRSV or **(E-H)** PPD. A two-way ANOVA for repeated measures with *post-hoc* Dunnet’s test corrected for multiple comparisons compared to Day 0 was performed for the data analysis. * = P<0.05.

**Figure 4.**
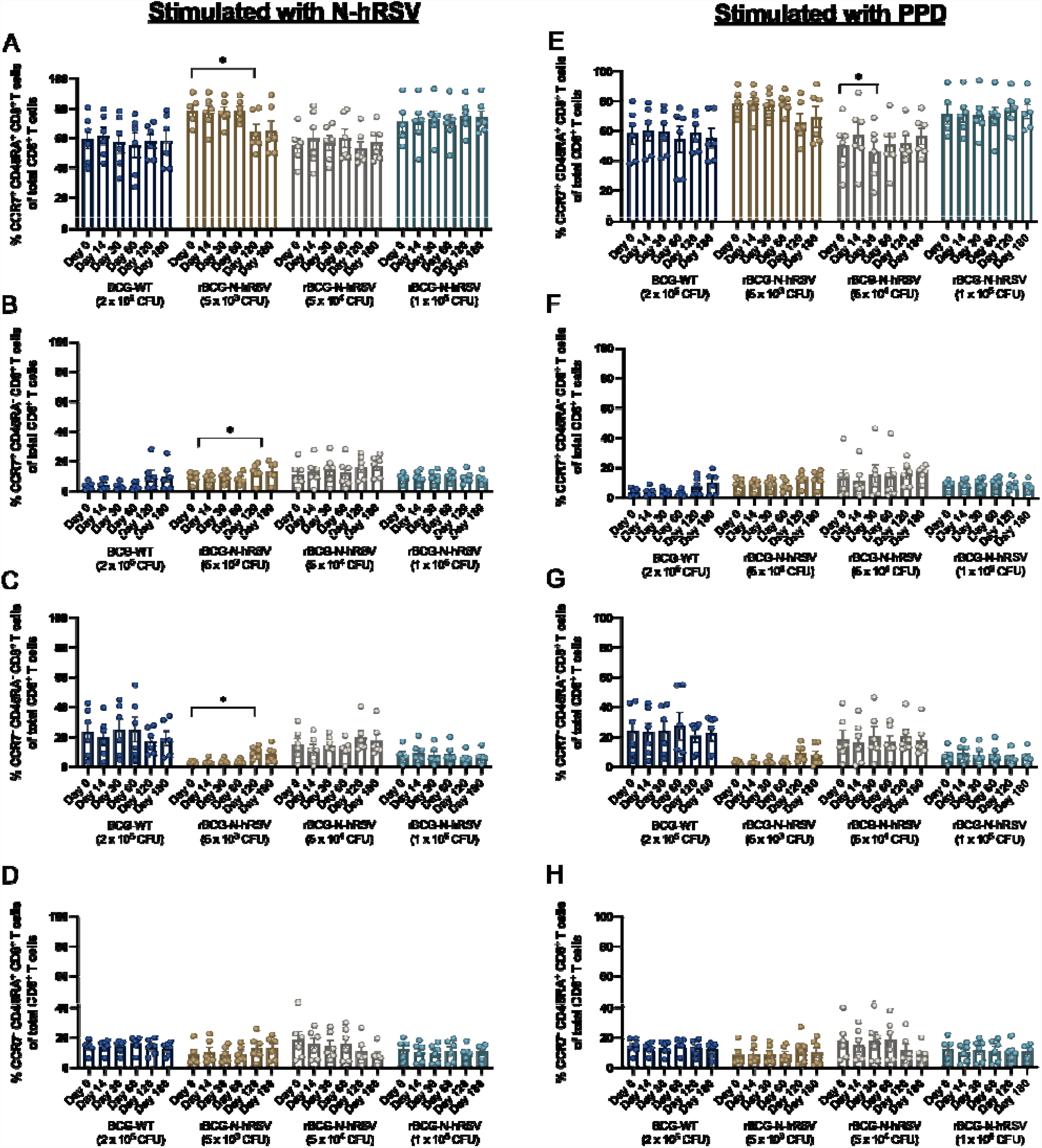
Quantification of CD8^+^ T cell memory subsets after a single immunization with rBCG-N-hRSV. T cell subsets defined by the expression of CCR7 and CD45RA were evaluated after stimulation of PBMCs with **(A-D)** N-hRSV or **(E-H)** PPD. Bars indicate means, while error bars represent SEM. A two-way ANOVA for repeated measures with *post-hoc* Dunnet’s test corrected for multiple comparisons compared to Day 0 was performed for the data analysis. * = P<0.05.

### PBMCs derived from rBCG-N-hRSV-vaccinated subjects produce cytokines in response to N-hRSV and mycobacterial antigens

Secretion of pro- and anti-inflammatory cytokines by PBMCs derived from rBCG-N-hRSV-vaccinated subjects in response to *in vitro* stimulation with N-hRSV (**Fig. 5**) or PPD (**Fig. 6**) was measured. Stimulation of PBMCs with N-hRSV led to increased levels of IFN-γ and IL-10 after immunization (**Fig. 5** and **Supp. Fig. 5**). Interestingly, the ratios of IL-6/IFN-γ and TNF-α/IFN-γ decreased in subjects immunized with 5×10^3^ or 5×10^4^ CFU of rBCG-N-hRSV (**Fig. 5G & H**). On the other hand, stimulation of PBMCs with PPD led to increased levels of IFN-γ, TNF-α, IL-2, and IL-6 (**Fig. 6** and **Supp. Fig. 6**) in subjects immunized with 5×10^3^ or 5×10^4^ CFU of rBCG-N-hRSV. Subjects immunized with 5×10^5^ CFU of BCG-WT also showed an upregulation of IL-6, as well as moderate IFN-γ, TNF-α, and IL-2 responses. A minor IL-10 response was detected for PBMCs from subjects immunized with 5×10^4^ CFU of rBCG-N-hRSV. These results suggest that immunization with either 5×10^3^ or 5×10^4^ CFU of rBCG-N-hRSV led to the secretion of pro-inflammatory cytokines typically associated with immune responses against intracellular pathogens, such as viruses and mycobacteria [16]–[19]. Moreover, the modest increase in IL-10 concentration is indicative of a balanced immune response [20].

**Figure 5.**
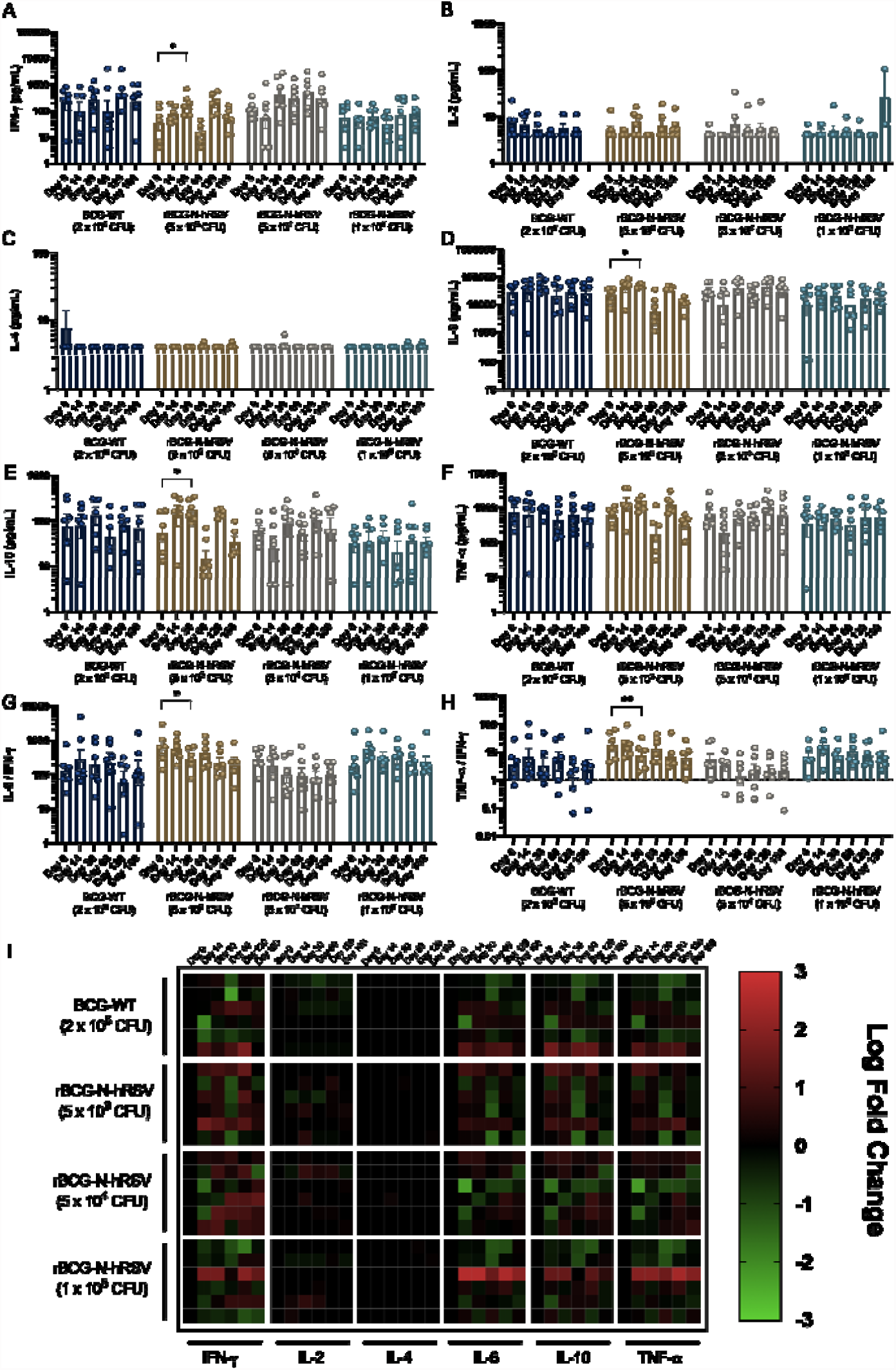
Stimulation with N-hRSV leads to IFN-γ and IL-10 secretion by PBMCs from subjects immunized with 5×10^3^ or 5×10^4^ CFU of rBCG-N-hRSV. The concentration of **(A)** IFN-γ, **(B)** IL-2, **(C)** IL-4, **(D)** IL-6, **(E)** IL-10, and **(F)** TNF-α in supernatants of PBMC cultures stimulated with 1.25 µg/mL N-hRSV are presented. Ratios between cytokines **(G)** IL-6 / IFN-γ and **(H)** TNF-α / IFN-γ were also calculated. Bars represent means and error bars represent SEM. A two-way ANOVA for repeated measures with *post-hoc* Dunnet’s test corrected for multiple comparisons compared to Day 0 over the log_10_ was performed for the analysis of the data. * = P<0.05, ** = P<0.01. **(I)** Heatmap of log_10_ fold change of the concentration of selected cytokines compared to Day 0. Each block of columns represents a particular cytokine, labeled below. Individual columns represent timepoints after immunization, specified above. Each block of rows represents a particular cohort of immunized study subjects, labeled left. Individual rows represent individual subjects. A color scale is depicted on the right.

**Figure 6.**
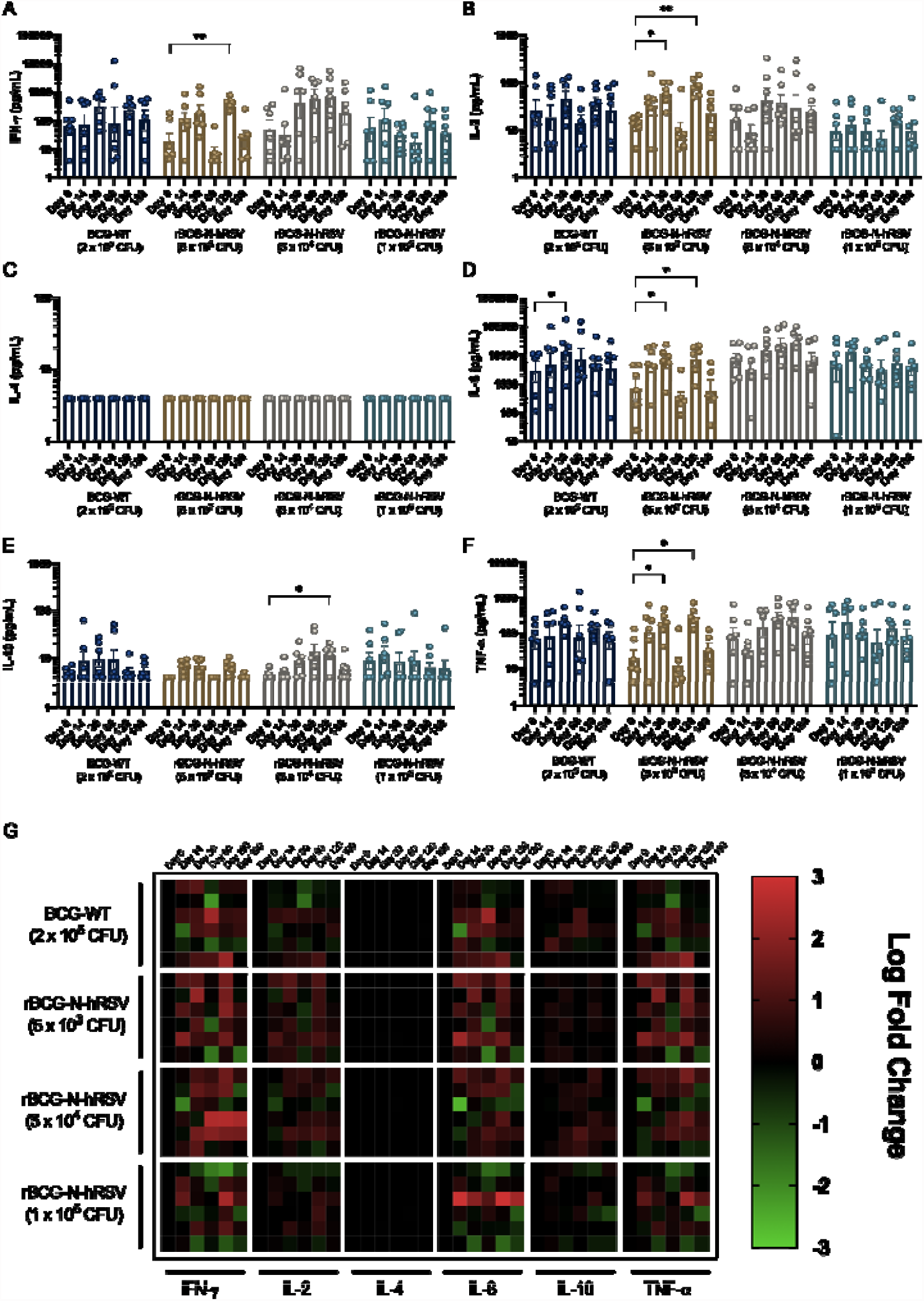
Stimulation with PPD leads to IFN-γ, TNF-α, and IL-6 secretion by PBMCs subjects immunized with 5×10^3^ or 5×10^4^ CFU of rBCG-N-hRSV. The concentration of **(A)** IFN-γ, **(B)** IL-2, **(C)** IL-4, **(D)** IL-6, **(E)** IL-10, and **(F)** TNF-α in supernatants of PBMC cultures stimulated with 750 IU/mL PPD are presented. Bars represent means and error bars represent SEM. A two-way ANOVA for repeated measures with *post-hoc* Dunnet’s test corrected for multiple comparisons compared to Day 0 over the log_10_ was performed for the analysis of the data. * = P<0.05, ** = P<0.01. **(G)** Heatmap of log_10_ fold change of the concentration of selected cytokines compared to Day 0. Each block of columns represents a particular cytokine, labeled below. Individual columns represent timepoints after immunization, specified above. Each block of rows represents a particular cohort of immunized study subjects, labeled left. Individual rows represent individual subjects. A color scale is depicted on the right.

### Reduced C1q binding by virus- and mycobacteria-specific antibodies induced by rBCG-N-hRSV

While it had been previously reported that the concentration of anti-N-hRSV antibodies in sera do not significantly change after immunization with rBCG-N-hRSV [7], it is possible that immunization with rBCG-N-hRSV may change the relative proportions of anti-N-hRSV IgG subclasses. Thus, the ability of anti-N-hRSV antibodies to bind exogenous C1q efficiently was measured as a functional assay for relative subclass composition of anti-N-hRSV IgG [21]. Exogenous C1q binding to anti-N-hRSV or anti-PPD antibodies in the sera was quantified for each sample (**Fig. 7A-B**). Both, anti-N-hRSV and anti-PPD antibodies from subjects immunized with 5×10^4^ CFU of rBCG-N-hRSV were found to bind less complement after immunization, as compared to the other groups (**Fig. 7A-B**). Importantly, these tendencies were maintained when C1q binding was normalized by total anti-N or anti-PPD IgG titers (C1q Binding Index) (**Fig. 7C-D**). Analyses of fold changes compared to the pre-immune condition showed similar results (**Supp. Fig. 7**).

**Figure 7.**
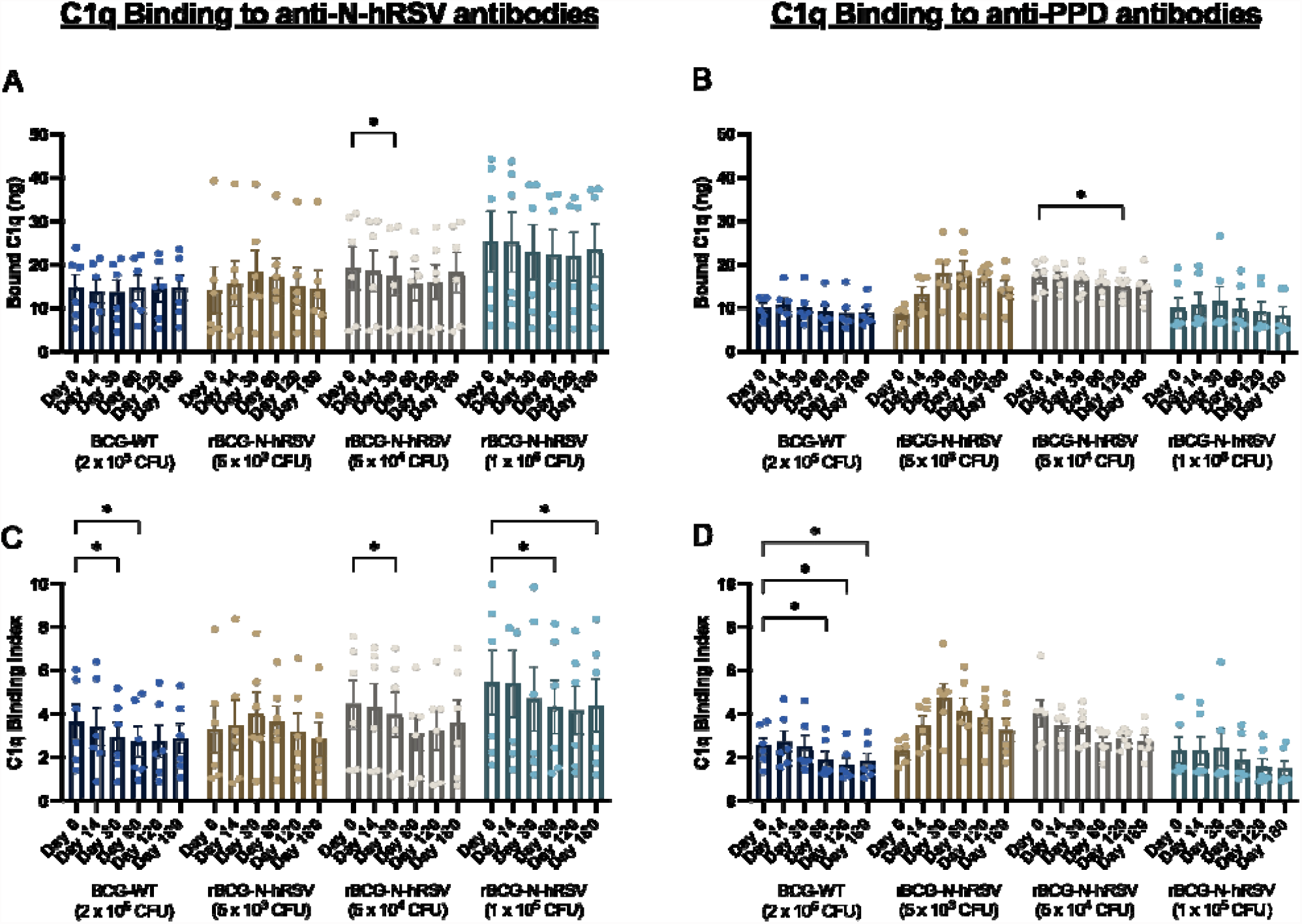
Reduced C1q binding by virus- and mycobacteria-specific antibodies induced by rBCG-N-hRSV. Total C1q binding is shown for **(A)** total anti-N-hRSV antibodies and **(B)** total anti-PPD antibodies. A C1q Binding Index was calculated by dividing total bound C1q by the log_10_ of total IgG against **(C)** N-hRSV or **(D)** PPD. Bars indicate means, while error bars represent SEM. A two-way ANOVA for repeated measures with *post-hoc* Dunnet’s test corrected for multiple comparisons compared to Day 0 was performed for the analysis of the data. * = P<0.05.

### rBCG-N-hRSV immunization induces equivalent IgG subclass secretion

The concentration of anti-N-hRSV and anti-PPD IgG1 and IgG2 in the sera of immunized subjects were quantified by ELISA. Immunization with rBCG-N-hRSV did not appear to change the concentration of these IgG subclasses, neither for anti-N-hRSV nor for anti-PPD antibodies (**Fig. 8**). As expected, IgG1 titers for anti-N-hRSV were higher than IgG2 titers (**Fig. 8A & C**). Surprisingly, IgG2 titers were higher than IgG1 titers for anti-PPD antibodies (**Fig. 8B & D**). Analyses of fold changes relative to the pre-immune condition showed similar results (**Supp. Fig. 8**).

**Figure 8.**
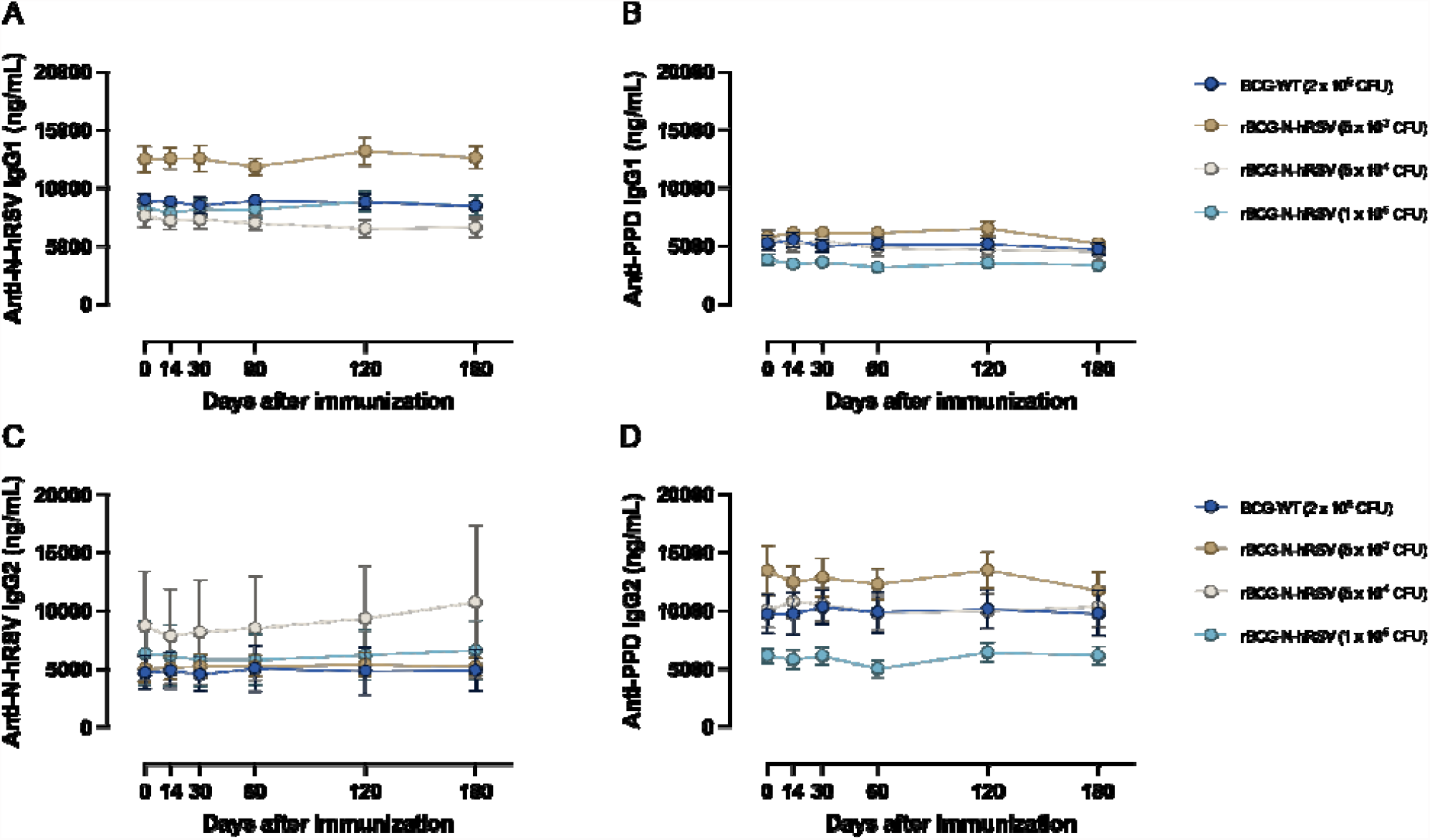
rBCG-N-hRSV immunization induces equivalent IgG subclass secretion. **(A-B)** IgG1 and **(C-D)** IgG2 antibody titers were measured via ELISA. Depicted are **(A & C**) anti-N-hRSV and **(B & D)** anti-PPD antibod concentrations over time. Dots represent mean titers, while error bars represent SEM. A two-way ANOVA for repeated measures with *post-hoc* Dunnet’s test corrected for multiple comparisons compared to Day 0 was performed for the analysis of the data.

## DISCUSSION

The results reported herein regarding cytokine concentrations in the sera constitute a good immunological indicator for the safety of rBCG-N-hRSV, given the moderate levels of circuliting cytokines detected (**Fig. 1**). A slight tendency of increased IFN-γ was observed for all experimental groups, except for those subjects immunized with the lowest dose of rBCG-N-hRSV. The increase in IFN-γ concentration can be interpreted as a positive result because this cytokine can be beneficial against hRSV [22]–[24]. This response is short-lived, may be dose-dependent [12], [13], [25] and is consistent with the notion that BCG can induce T_H_1 responses in humans [26]–[28], which is one of the reasons as to why it was selected as a vector for immunizing against hRSV antigens [7]–[11], [13], [29], [30].

Furthermore, rBCG-N-hRSV had previously been shown to be an efficient inducer of T_H_1 cellular responses against N-hRSV in healthy adults in this clinical trial, as measured by the secretion of IFN-γ by PBMCs [7]. Our results further support this notion since subjects immunized with 5×10^4^ CFU of rBCG-N-hRSV showed a considerable Perforin and Granzyme B response against PPD and N-hRSV (**Fig. 2**), as well as by the increase in IFN-γ concentration in cultures of stimulated PBMCs (**Figs. 5 & 6**). The heterogenous secretion of cytotoxic molecules shown by PBMCs from subjects immunized with different doses of rBCG-N-hRSV in response to N-hRSV could be explained by differential exposure to community hRSV during seasonal outbreaks by this virus.

Even though we detected no expansion of memory T cells in response to immunization with either BCG-WT or rBCG-N-hRSV alone (**Figs. 3 & 4**), natural infection with hRSV after immunization with rBCG-N-hRSV could enhance the expansion of memory T cell subsets, as seen for other viruses [31], [32]. However, this remains to be determined, as none of the study subjects reported hRSV-associated symptoms during the study.

Lastly, decreased complement binding via the classical pathway was observed after immunization with rBCG-N-hRSV, especially at the higher doses (**Fig. 7**). However, no detectable changes in IgG1 or IgG2 concentrations were found for antibodies specific against N-hRSV or PPD (**Fig. 8**). The relevance of this subtle change in antibody effector function remains to be determined, but it seems a good indicator of the safety for this vaccine candidate [33].

Limitations of this study include the low sample size, considering that the primary outcome of the clinical trial was to evaluate safety for this recombinant BCG vaccine. Moreover, the immune response evaluated herein was assessed only during an immune steady state, so differential immune responses after infection are yet to be determined. Lastly, while assessing the immune response elicited by rBCG-N-hRSV in healthy male adults was necessary before evaluating this vaccine in the pediatric population, the response elicited in newborns and infants would be particularly relevant, considering that they are the target population for immunization against hRSV. Our data support the immunological safety and appropriateness of rBCG-N-hRSV as a vaccine against this viral pathogen, upon further clinical efficacy evaluation.

## Data Availability

All data produced in the present study are available upon reasonable request to the authors.

## AUTHOR CONTRIBUTIONS

Conceptualization: G.A.P., N.M.S.G., C.A.A., P.A.G., S.M.B., A.M.K.

Visualization: G.A.P., N.M.S.G., C.A.A., P.A.G., S.M.B., A.M.K.

Methodology: G.A.P., N.M.S.G., C.A.A., L.R-G., Y.V.

Investigation: G.A.P., N.M.S.G., C.A.A., L.R-G., Y.V., P.A.G., S.M.B., A.M.K.

Funding acquisition: P.A.G., S.M.B., A.M.K.

Project administration: P.A.G., S.M.B., A.M.K.

Supervision: P.A.G., S.M.B., A.M.K.

Writing – original draft: G.A.P., A.M.K.

Writing – Review and editing: G.A.P., N.M.S.G., C.A.A., L.R-G., Y.V., P.A.G., S.M.B., A.M.K.

