## Supplementary Figures for "A recombinant BCG-based vaccine against the human respiratory syncytial virus induces a balanced cellular immune response against viral and mycobacterial antigens"

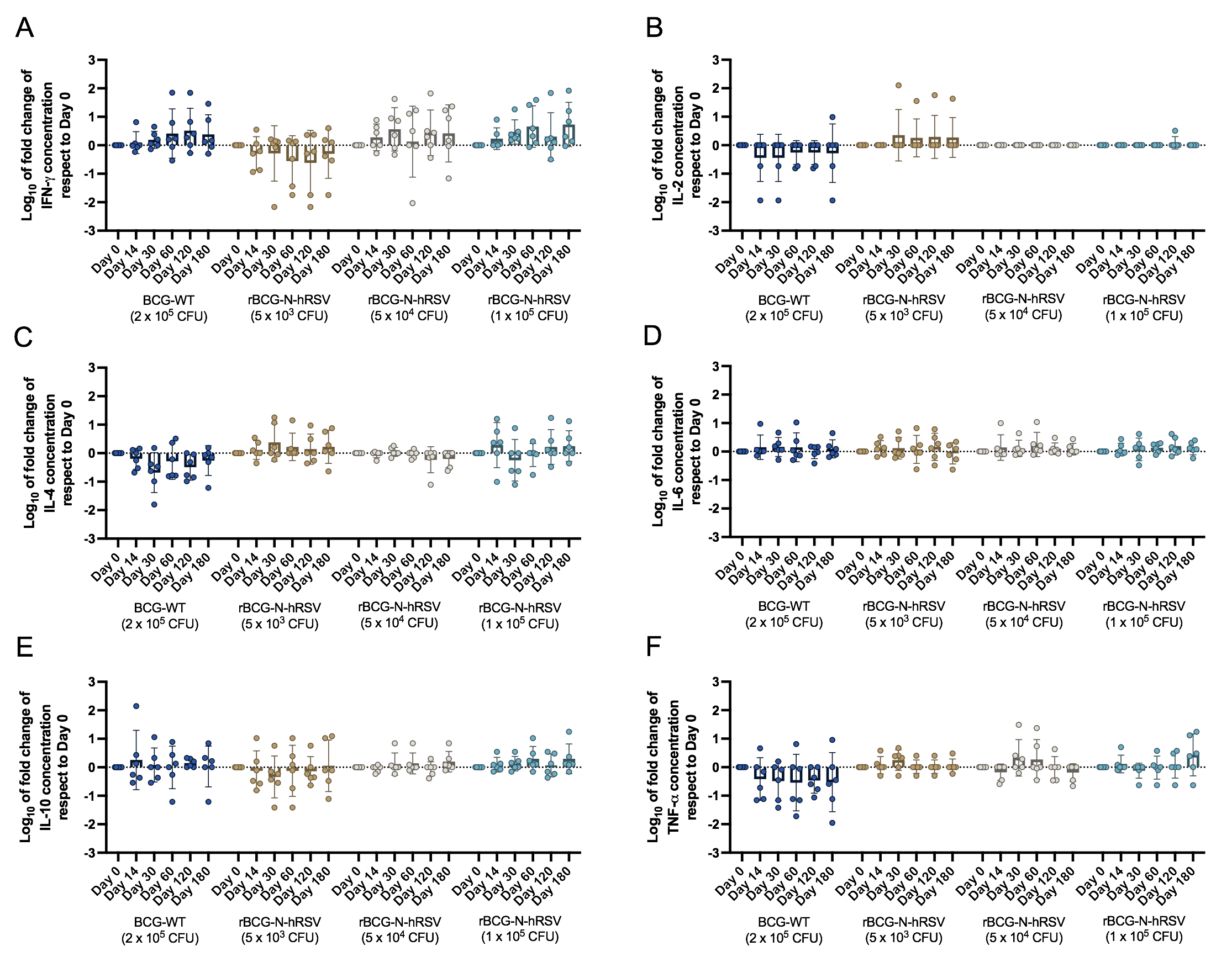

**Supplementary Figure 1. Fold changes in cytokine concentrations in sera of immunized subjects.** Base 10 logarithms of fold changes for the concentration of **(A)** IFN-γ, **(B)** IL-2, **(C)** IL-4, **(D)** IL-6, **(E)** IL-10, and **(F)** TNF-α in sera samples as compared to day 0 are presented. Bars represent means and error bars represent SEM. A two-way ANOVA for repeated measures with *post-hoc* Dunnet’s test corrected for multiple comparisons relative to Day 0 was performed for the analysis of the data.

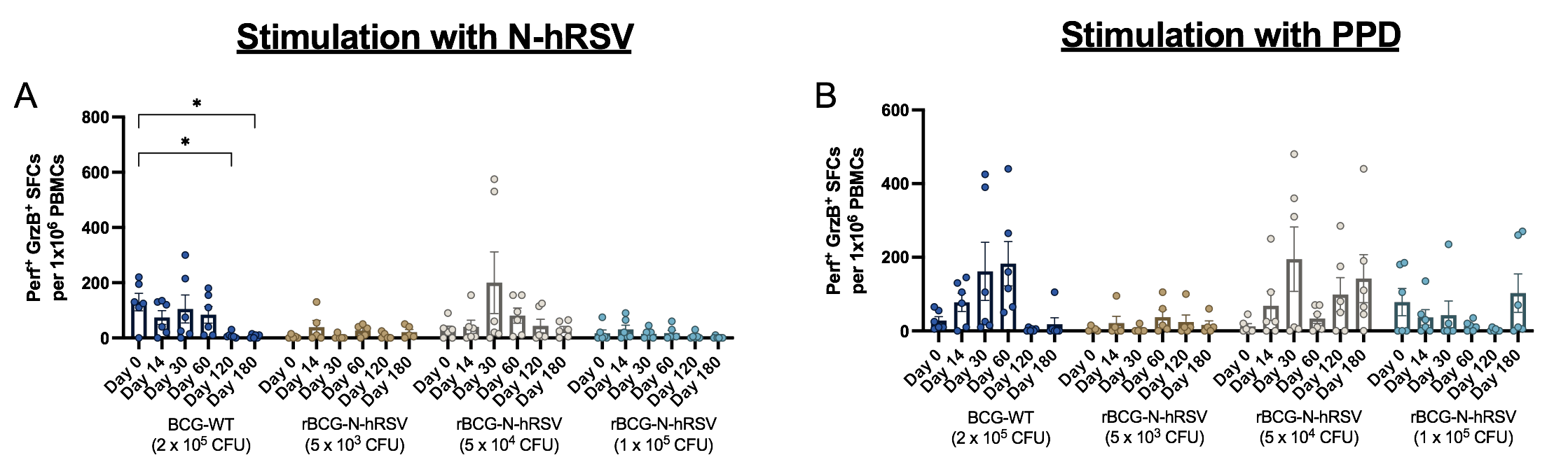

**Supplementary Figure 2. Double producers of Perforin and Granzyme B measured by ELISPOT assays.** Perf^+^ GrzB^+^ spot forming cells (SFCs) were counted after PBMCs were stimulated for 48 hours with either 1.25 µg/mL N-hRSV **(A)** or 750 IU/mL PPD **(B)**. Bars represent the mean value of SFCs, and error bars represent the SEM. A two-way ANOVA for repeated measures with *post-hoc* Dunnet’s test corrected for multiple comparisons relative to Day 0 was performed for the analysis of data. * = P<0.05.

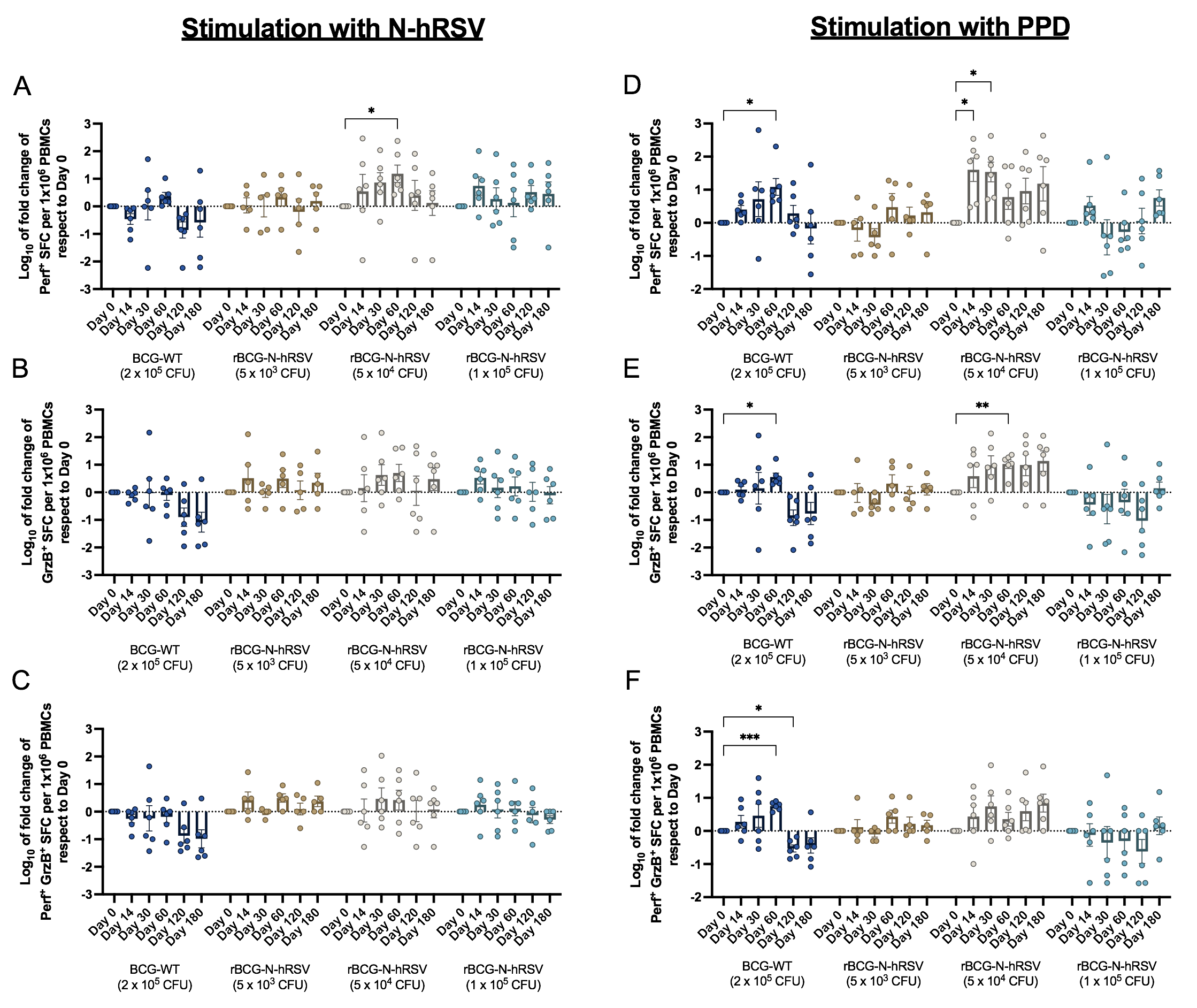

**Supplementary Figure 3. Fold changes of Perforin^+^ and Granzyme B^+^ spot forming cells.** Base 10 logarithms of fold changes of **(A, D)** Perf^+^, **(B, E)** GrzB^+^, or **(C, F)** Perf^+^ GrzB^+^ SFCs as compared to day 0 are shown. PBMCs were stimulated for 48 hours with either 1.25 µg/mL N-hRSV **(A-C)** or 750 IU/mL PPD **(D-F)**. Bars represent the mean value of SFCs, and error bars represent the SEM. A two-way ANOVA for repeated measures with *post-hoc* Dunnet’s test corrected for multiple comparisons relative to Day 0 was performed for the analysis of data. * = P<0.05, ** = P<0.01, *** = P<0.005.

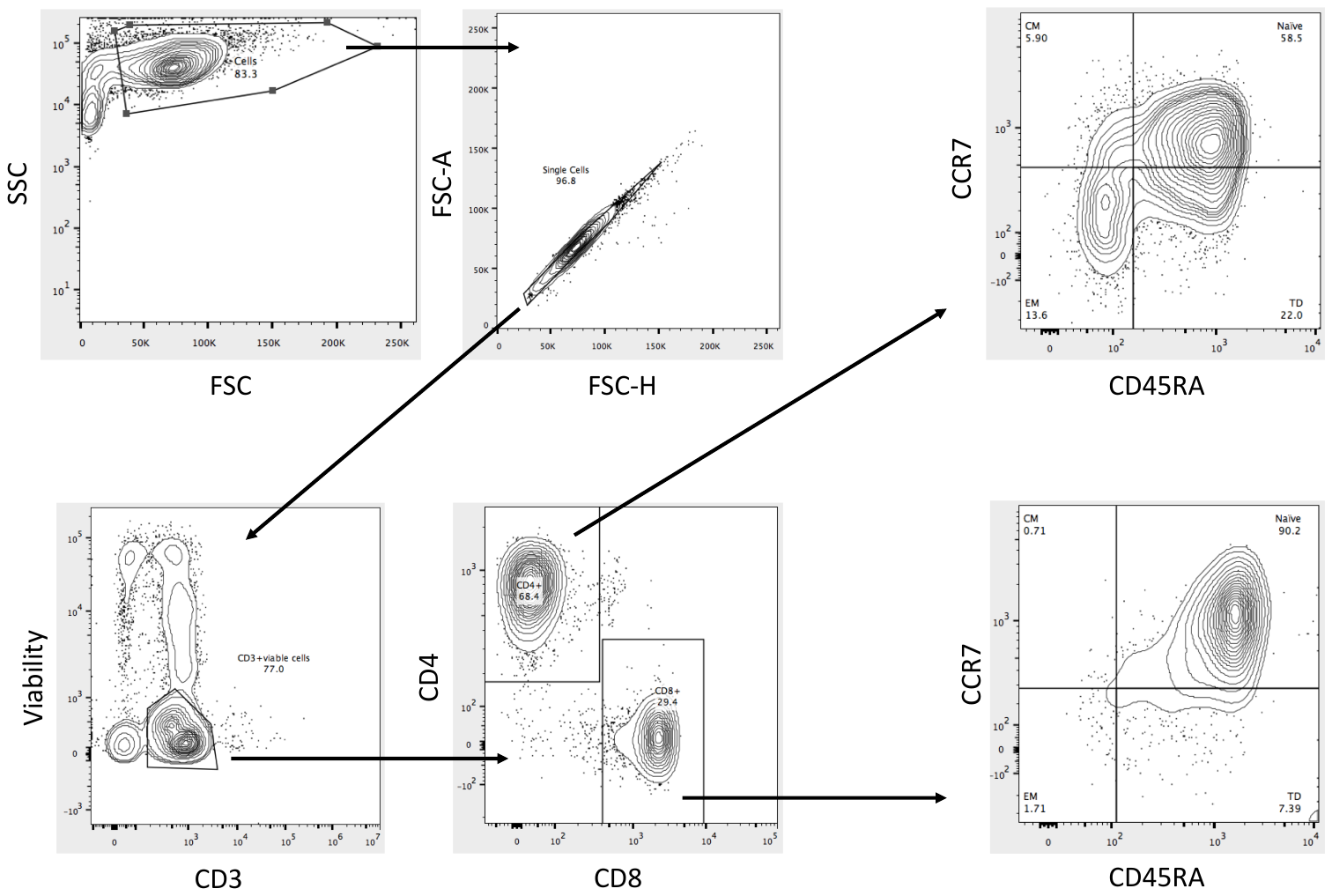

**Supplementary Figure 4. Gating strategy for the identification of T cell memory subsets.** T cell subsets defined by the expression of CCR7 and CD45RA were evaluated after stimulation of PBMCs with N-hRSV or PPD. CD4^+^ or CD8^+^ T cell populations were identified as naïve (CCR7^+^ CD45RA^+^), central memory (T_CM_, CCR7^+^ CD45RA^-^), effector memory (T_EM_, CCR7^-^ CD45RA^-^), or CD45RA-expressing effector memory (T_EMRA_, CCR7^-^ CD45RA^+^).

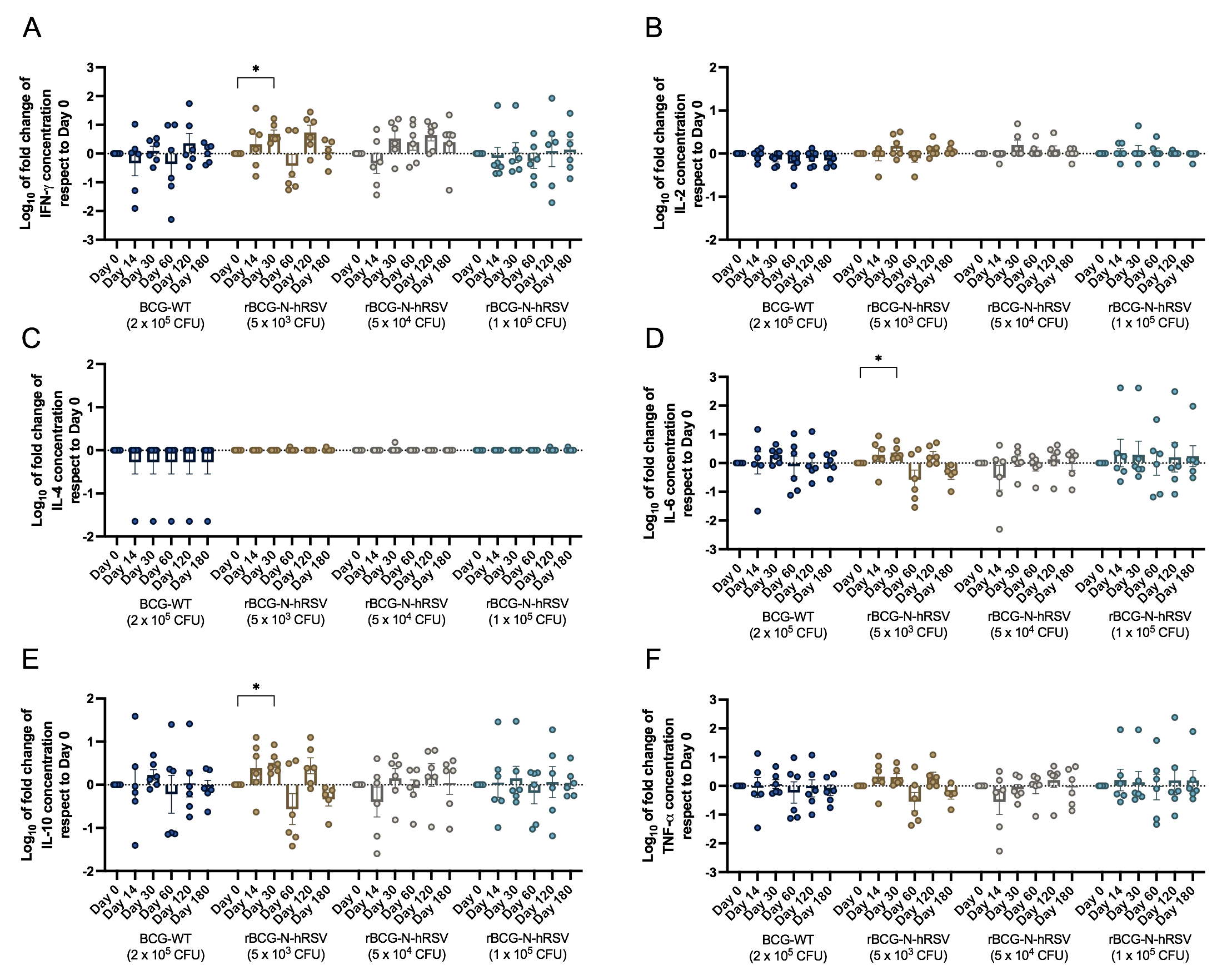

**Supplementary Figure 5. Fold changes in cytokine concentrations of supernatants of PBMC cultures stimulated with N-hRSV.** Base 10 logarithms of fold changes of concentration of **(A)** IFN-γ, **(B)** IL-2, **(C)** IL-4, **(D)** IL-6, **(E)** IL-10, and **(F)** TNF-α in supernatants of PBMC cultures stimulated with 1.25 µg/mL N-hRSV are presented. Bars represent means and error bars represent SEM. A two-way ANOVA for repeated measures with *post-hoc* Dunnet’s test corrected for multiple comparisons relative to Day 0 over the base 10 logarithm was performed for the analysis of the data. * = P<0.05.

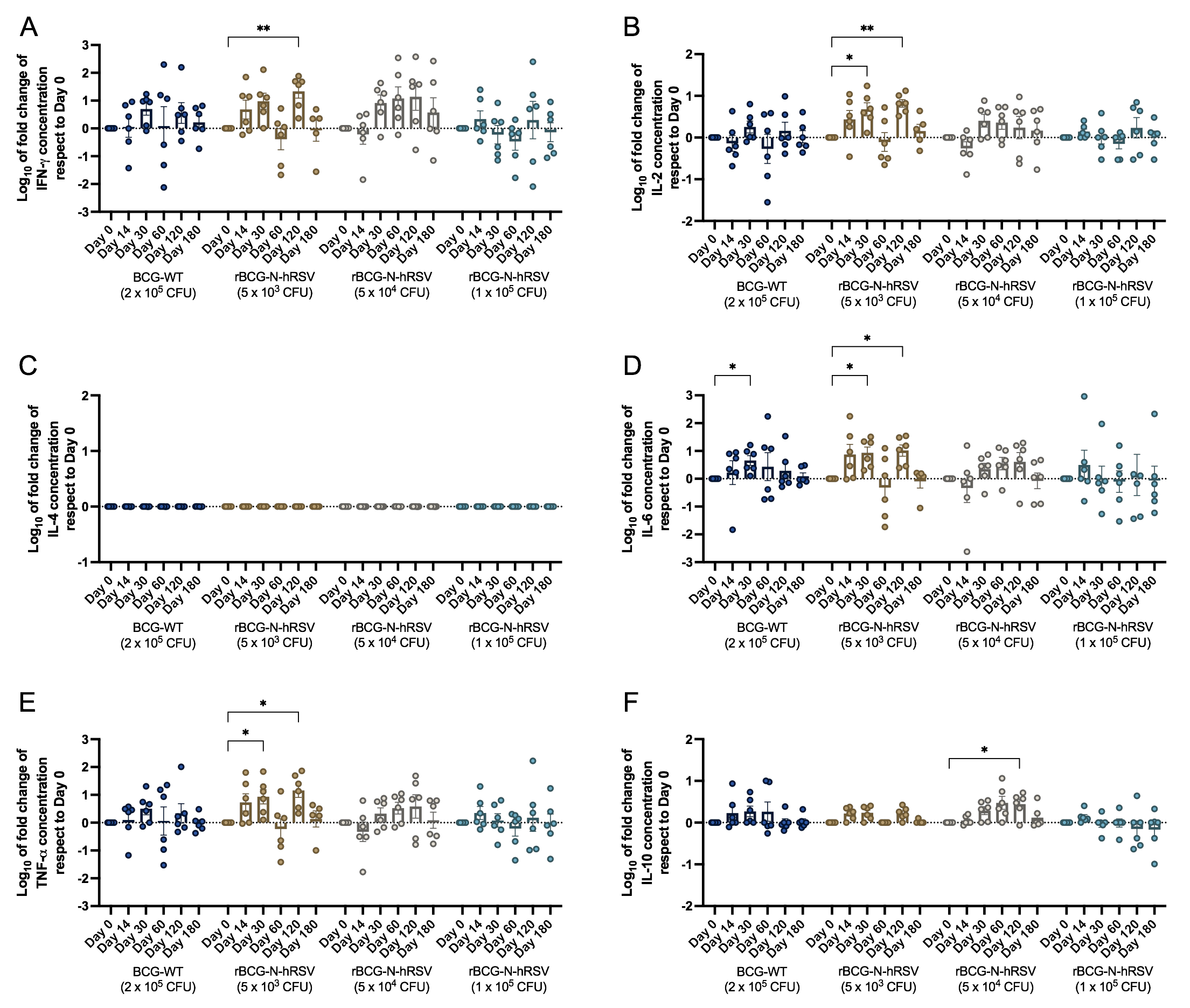

**Supplementary Figure 6. Fold changes in cytokine concentrations of supernatants of PBMC cultures stimulated with PPD.** Base 10 logarithms of fold changes of concentration of **(A)** IFN-γ, **(B)** IL-2, **(C)** IL-4, **(D)** IL-6, **(E)** IL-10, and **(F)** TNF-α in supernatants of PBMC cultures stimulated with 750 IU/mL PPD are presented. Bars represent means and error bars represent SEM. A two-way ANOVA for repeated measures with *post-hoc* Dunnet’s test corrected for multiple comparisons relative to Day 0 over the base 10 logarithm was performed for analysis of the data. * = P<0.05, ** = P<0.01.

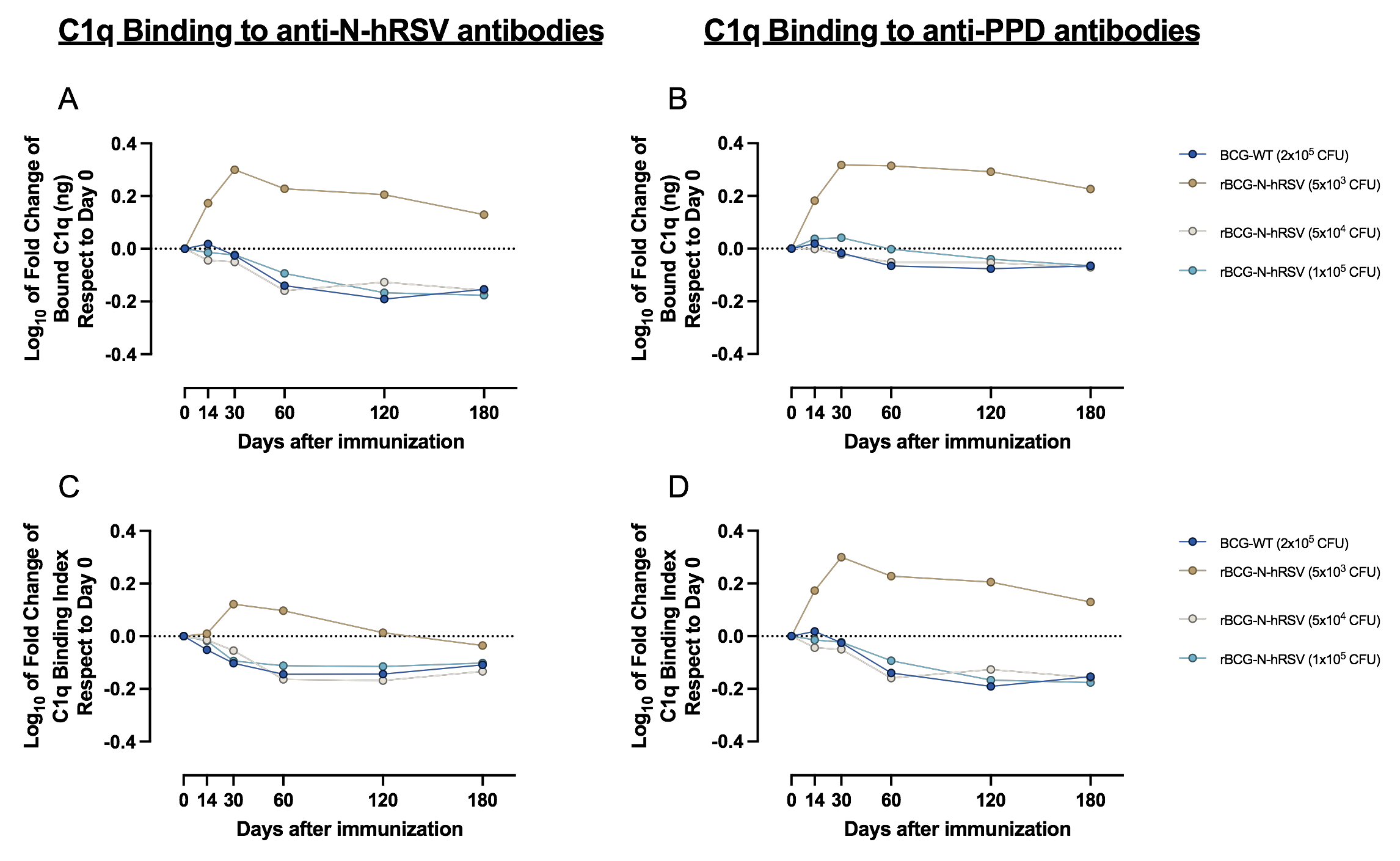

**Supplementary Figure 7. Fold changes in C1q binding by antibodies elicited by immunization.** Base 10 logarithms of fold changes of **(A, B)** total C1q binding and **(C, D)** C1q binding index is shown for **(A, C)** total anti-N-hRSV antibodies and **(B, D)** total anti-PPD antibodies. Bars indicate means. A two-way ANOVA for repeated measures with *post-hoc* Dunnet’s test corrected for multiple comparisons relative to Day 0 was performed for analysis of the data.

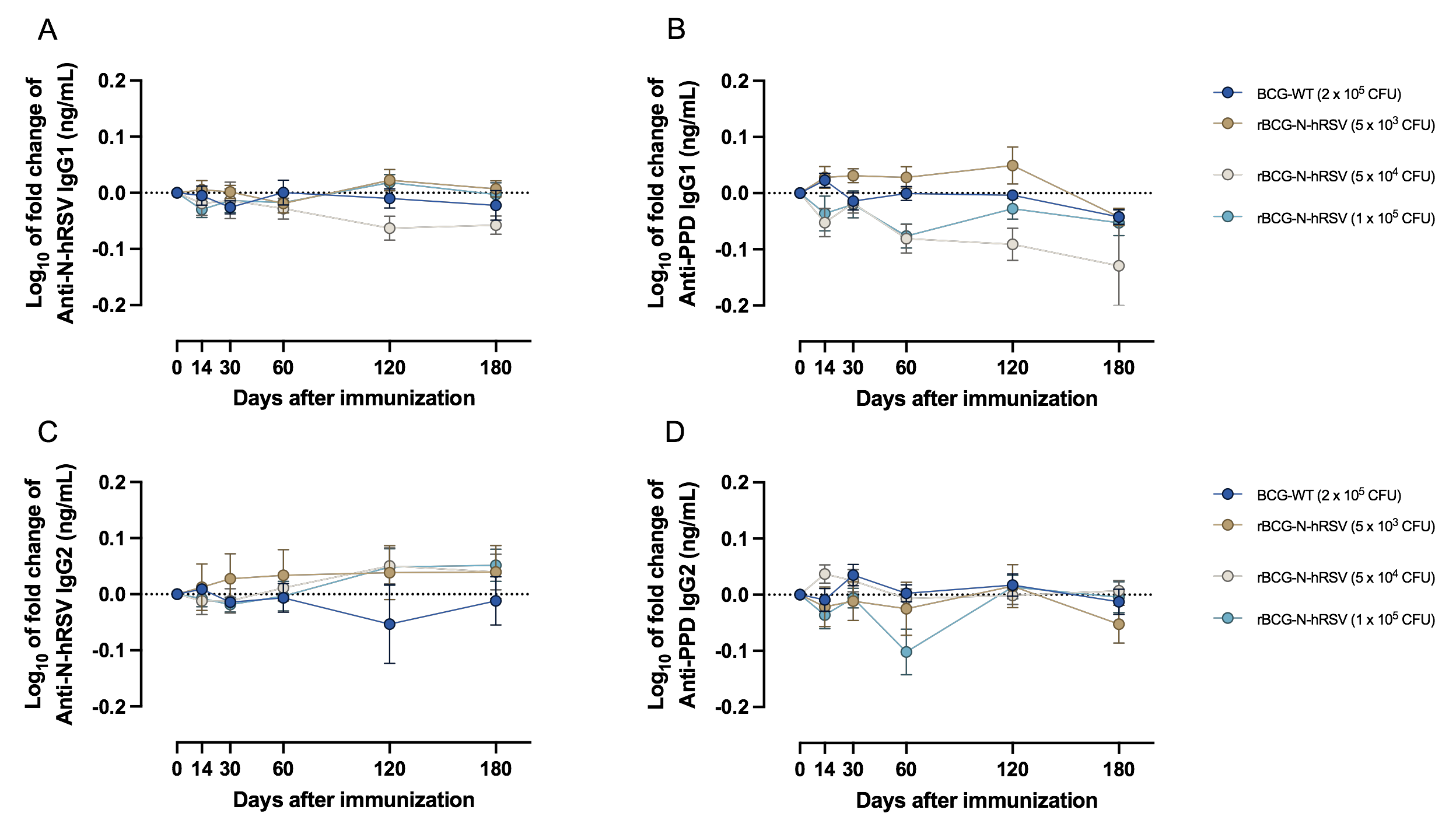

**Supplementary Figure 8. Fold changes in IgG1 and IgG2 antibodies elicited by immunization.** Base 10 logarithms of fold changes of **(A, B)** anti-N-hRSV antibodies and **(C, D)** anti-PPD antibodies is shown for **(A, C)** IgG1 and **(B, D)** IgG2 antibodies. Bars indicate means and error bars represent SEM. A two-way ANOVA for repeated measures with *post-hoc* Dunnet’s test corrected for multiple comparisons relative to Day 0 was performed for analysis of the data.
